# Knowledge-guided Deep Temporal Clustering for Alzheimer’s Disease Subtypes in Completed Clinical Trials

**DOI:** 10.1101/2023.10.13.23296985

**Authors:** Dulin Wang, Xiaotian Ma, Paul E. Schulz, Xiaoqian Jiang, Yejin Kim

## Abstract

Alzheimer’s disease (AD) is a multifaceted neurodegenerative disorder with varied patient progression. We aim to test the hypothesis that AD patients can be categorized into subgroups based on differences in progression. We leveraged data from three randomized clinical trials (RCTs) to develop a knowledge-guided, deep temporal clustering (KG-DTC) framework for AD subtyping. This model combined autoencoders for contextual information capture, k-means clustering for representation formation, and clinical outcome classification for clinical knowledge integration. The derived representations, encompassing demographics, APOE genotype, cognitive assessments, brain volumes, and biomarkers, were clustered using the Gaussian Mixture Model to identify AD subtypes. Our novel KG-DTC framework was developed using placebo data from 2,087 AD patients across three solanezumab clinical trials (EXPEDITION, EXPEDITION2, and EXPEDITION3), achieving high performance in outcome prediction and clustering. The KG-DTC model demonstrated superior clustering structures, especially when combined with k-means clustering loss. External validation with independent clinical trial data showed consistent clustering results, with a 0.33 silhouette score for three clusters. The model’s stability was confirmed through a leave-one-out approach, with an average adjusted Rand Index around 0.945. Three distinct AD subtypes were identified, each exhibiting unique patterns of cognitive function, neurodegeneration, and amyloid beta levels. Notably, Subtype 3 (S3) showed rapid cognitive decline across multiple clinical measures (e.g., 0.64 in S1 vs. −1.06 in S2 vs. 15.09 in S3 of average ADAS total change score, p<.001). This innovative approach offers promising insights for understanding variability in treatment outcomes and personalizing AD treatment strategies.

## Introduction

Alzheimer’s disease (AD) is incurable and challenging to diagnose and treat due to its complexity, heterogeneity, and multifactor nature. Its progression patterns manifest through a broad spectrum of longitudinally linked clinical features and outcomes that vary across AD patients. Variability in AD has impeded many clinical trials for drug development^1,2^. Therefore, precision medicine for AD aims to understand why individuals respond differently to treatments, and temporal subtyping has become a crucial tool in identifying patient subgroups to answer such questions. By transforming the raw data along disease progression into clinically relevant and interpretable information, temporal subtyping minimizes progression heterogeneity among individuals^3,4^, guides clinicians to tailor treatment to AD subgroups, and ultimately develops successful AD subtype-specific drugs.

Recent studies have used electronic health records (EHRs) and Alzheimer’s Disease Neuroimaging Initiative (ADNI) databases extensively to derive systematic and comprehensive AD subtypes^5–8^. However, these datasets are sparse or noisy. Randomized clinical trials (RCTs) are another rich but understudied multimodal data source. The placebo groups from RCTs allow us to investigate the disease progression without being confounded by the exposure to experimental drugs. A pooled group of placebo-treated patients from RCTs can increase the power and diversity of AD populations. RCT databases were used to identify phenotypes or subtypes in sepsis and respiratory disease^9,10^. Considering the successful utilization of RCT data in various diseases, using multimodal data from RCT to derive the AD subtypes is promising.

Temporal subtyping of AD is a data-driven, unsupervised learning task to group patients into similar disease progression patterns. Clustering is a widely used to discover subtype. Recent advances in deep representation learning overcome the limitations of traditional clustering methods that are hard to handle high-dimensional and multimodal data^6,11^. The deep learning-based clustering learns low-dimensional representation for multivariate longitudinal observations, which can be used in downstream tasks. However, the separate step learning approach has certain limitations. Firstly, it learns a low-dimensional representation for multivariate longitudinal observations, but this is not directly learned for clustering, which can hinder the overall clustering performance. Secondly, this method identifies temporal subtypes in an entirely unsupervised manner, ignoring any existing information about patients’ clinical outcomes. This information, such as clinical trial outcomes, is crucial for understanding the progression of the disease and predicting future clinical outcomes. Thus, such an approach may not fully utilize all available resources for optimal results.

This study proposed knowledge-guided, deep temporal clustering, which is a unified framework to identify AD subtypes. We first developed a knowledge-guided deep clustering architecture to derive clustering-friendly vector-based representations. This architecture combined (1) temporal autoencoders (AEs) to capture contextual information and generate representations, (2) *k*-means clustering to encourage the representation to form clusters, and (3) clinical outcome classification to reflect clinical knowledge. Second, our model could generate informative representation through qualitative and quantitative analysis to derive meaningful subtypes. To this end, we applied the proposed framework to construct the representation learned from pooled RCTs with multimodal data, namely demographics, cognitive assessments, brain region volumes, biomarkers including amyloid-beta (*Aβ*), and genomic data on the apolipoprotein E (APOE) gene. We used the learned representation to cluster patients through the Gaussian mixture model (GMM) to derive AD subtypes and characterize the subtypes of AD to interpret potential disease progression patterns. From our extensive validation on the subtypes via reproducibility test, stability test, and significance test, we found that the subtypes were reproducible, stable, and clinically meaningful.

## Results

### Data summary

We developed our model using 2,087 AD patients from three RCT placebo arms (505 in EXPEDITION1^13^, 518 from EXPEDITION2^13^, and 1,064 from EXPEDITION3^14^). For each patient, we included visits with assessments conducted or biomarkers collected. As a result, we selected visits 1, 2, 3, 4, 5, 6, 10, 13, 16, 19, and 23.0 for EXPEDITION 1 and 2, and visits 1, 2, 3, 5, 9, 12, 15, 18, and 22 for EXPEDITION 3. The total number of visits was 13,946. We set aside EXPEDITION2 as an external validation set and used EXPEDITION1 and EXPEDITION3 as a discovery set. The data comprises eight demographics, 28 baseline efficacy measurements, and 28 time-variant variables observed from baseline to the end of clinical trials. The efficacy measures include cognitive assessments, imaging biomarkers, fluid biomarkers, quality-of-life assessments, and a neuropsychiatric assessment (Supplementary Table S1. Demographics and longitudinal features summary.).

### Knowledge-guided deep temporal clustering summary

We developed a novel and unified framework (Fig. 1) for knowledge-guided deep temporal clustering (KG-DTC, Fig. 5) to identify subtypes from temporal multimodal data (Method: Subtype discovery: Knowledge-Guided Deep Temporal Clustering (KG-DTC) model).

**Fig. 1.**
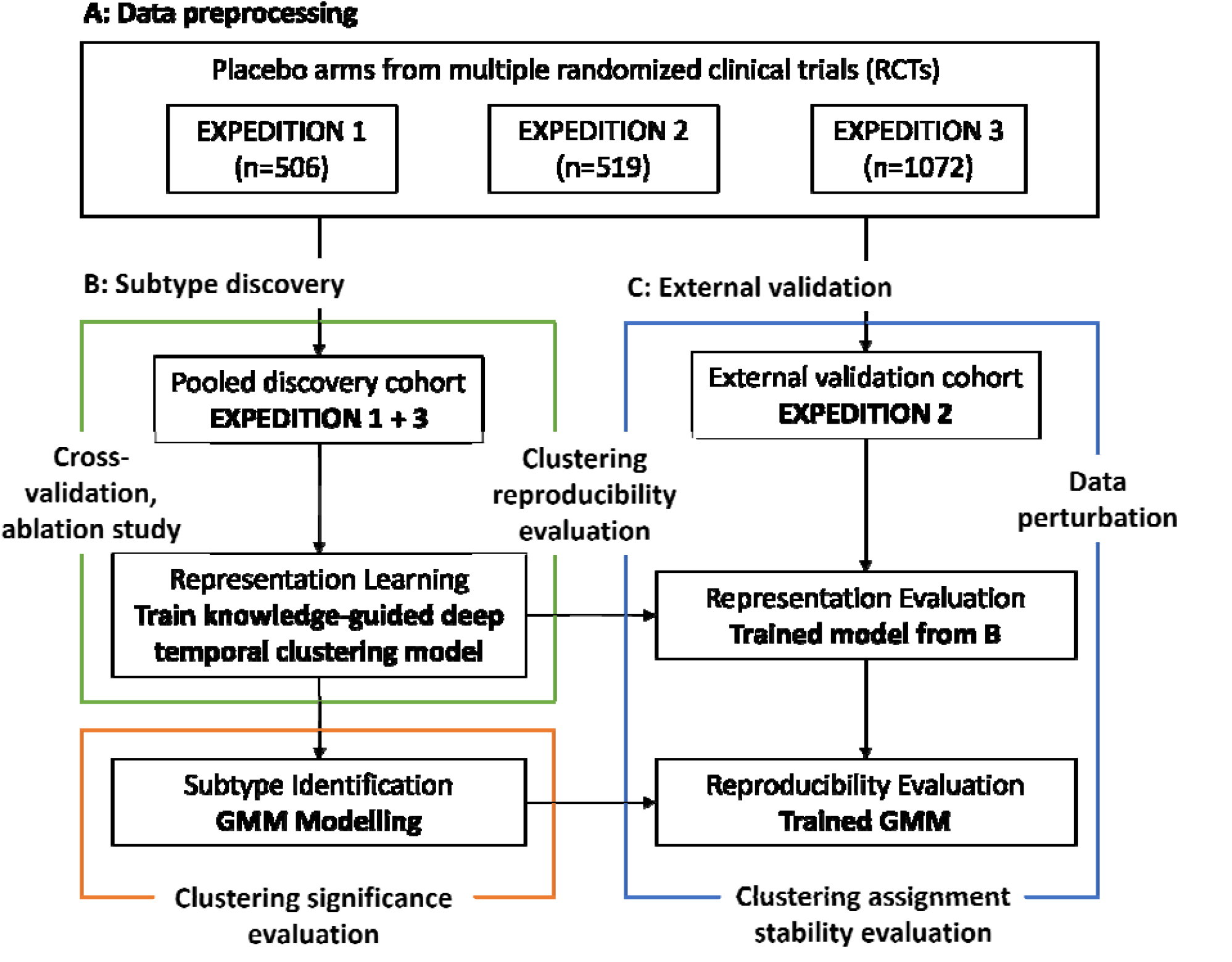
Overall framework. Our framework to identify the subtypes of AD progression has three stages: 1) data preprocessing, 2) subtype discovery, and 3) external validation. After data preprocessing and pooling, we developed knowledge-guided deep temporal clustering (KG-DTC methods and applied them to pooled clinical trial data. The KG-DTC method identified subtypes of AD patients using their longitudinal observations on efficacy measures. We internally validated the model using cross-validation and ablation study. We then externally validated the clustering result’s reproducibility and stability with independent clinical trial data. After thoroughly evaluating the clustering model, we investigated the characteristics of clusters (i.e., subtypes) and common patterns within the clusters. This figure was adapted from Dinga.^12^

### Model cross-validation

We first evaluated the performance of KG-DTC in a cross-validation manner. We evaluated whether our model could learn a representation that can predict clinical outcomes when embedding the knowledge guidance. We also evaluated our model’s clustering performance by silhouette scores and UMAP (uniform manifold approximation and projection for dimension reduction) visualization (Method: Model evaluation). For comparison, we ablated each component in KG-DTC (i.e., *k*-means clustering, knowledge guidance) and evaluated the three criterions above. Overall, we found that KG-DTC achieved high performance in both outcome prediction and clustering (Table 1). KG-DTC had *R*-squared (*R*^2^) scores of 0.84 and 0.31 for two selected clinical outcome variables (i.e., ADAS and CDR) and a silhouette score of 0.26 with three clusters. The UMAP plot showed that three clusters separate the patients well. Other ablated models (M1, M2, and M3) failed to achieve a balance between outcome prediction and clustering. The UMAP showed that the clusters are not compact or not evenly separated.

**Table 1.**
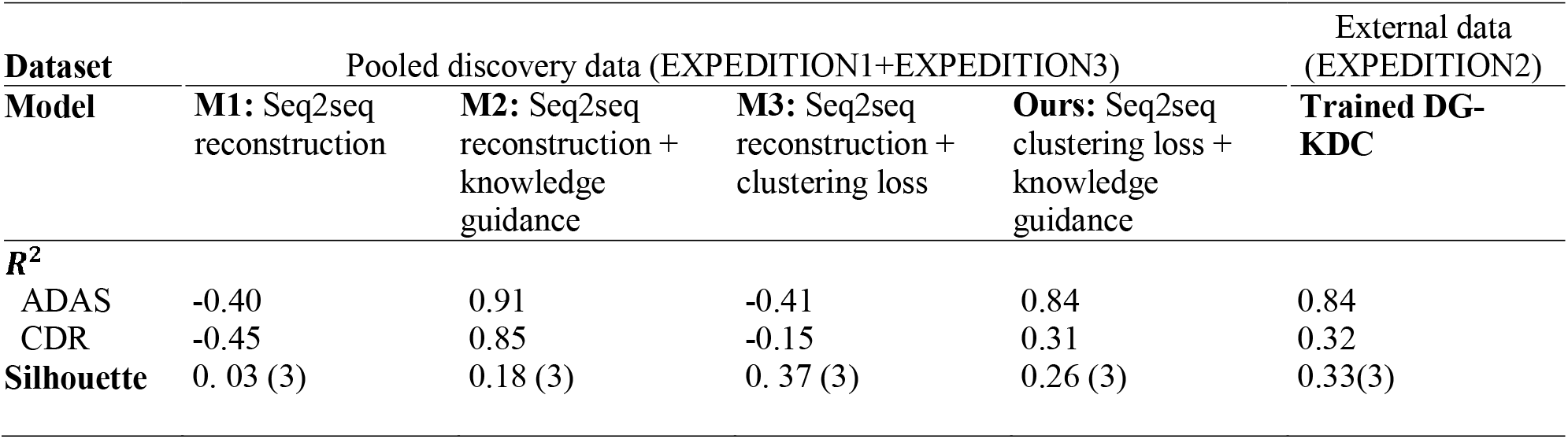
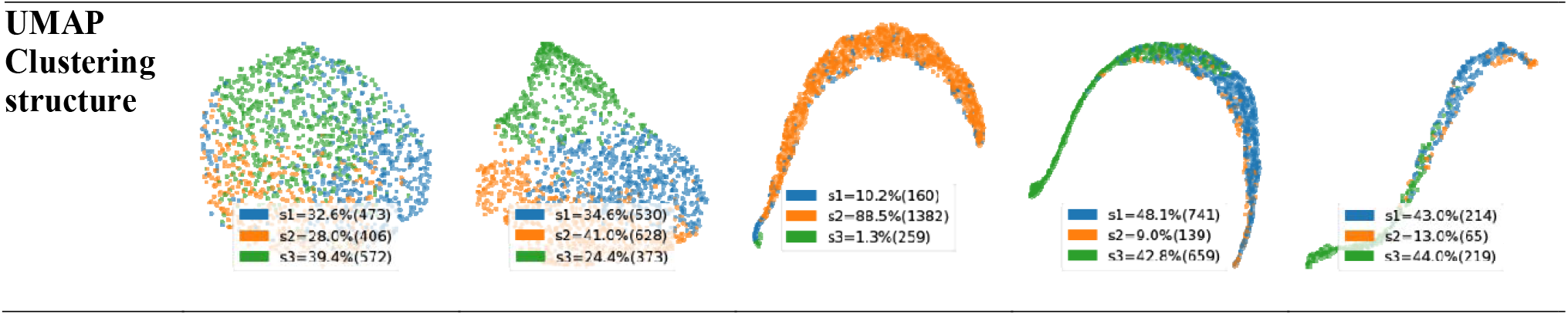
Evaluation of representation and clustering with ablated models, and reproducibility of clustering. Silhouette: Highest silhouette scores (number of clusters). We use scores to examine outcome regression fit to evaluate representation quality; we use the highest silhouette scores with the number of clusters for clustering performance evaluation; we use UMAP visualization for clustering structure evaluation. We evaluated the model’s representation, clustering, and effectiveness of components with three ablated models: 1) M1: simple model with only seq2seq reconstruction structure, 2) M2: model with seq2seq reconstruction and knowledge guidance, and 3) M3: model with seq2seq reconstruction and clustering loss.

| Dataset | Pooled discovery data (EXPEDITION1+EXPEDITION3) |  |  |  | External data (EXPEDITION2) |
| --- | --- | --- | --- | --- | --- |
| Model | M1: Seq2seq reconstruction | M2: Seq2seq reconstruction + knowledge guidance | M3: Seq2seq reconstruction + clustering loss | Ours: Seq2seq clustering loss + knowledge guidance | Trained DG-KDC |
| $R^2$ | | | | | |
| ADAS | -0.40 | 0.91 | -0.41 | 0.84 | 0.84 |
| CDR | -0.45 | 0.85 | -0.15 | 0.31 | 0.32 |
| Silhouette | 0.03 (3) | 0.18 (3) | 0.37 (3) | 0.26 (3) | 0.33(3) |

We then investigated the contribution of each model component. We observed that joint optimization of *k*-means clustering significantly improved silhouette scores and made compact clustering structures in UMAP visualization. As shown in Table 1. UMAP clustering structure, it was evident that the representations learned by M3 and KG-DTC with *k*-means clustering loss have formed clearly separated and compressed clusters. In contrast, the representations learned by the ablated models M1 and M2 without *k*-means clustering loss fail to form distinct cluster shapes (highest silhouette scores of 0.03 and 0.18 at cluster counts 3, respectively), and the samples are scattered with a great amount of mixing. In addition, we observed that knowledge guidance helps preserve rich clinical context in representation. Knowledge guidance increased the outcome prediction accuracy (M1’s of −0.40 and −0.45 vs. M3’s of −0.41 and −0.15), and it also prevented the accuracy from extremely decreasing when it was combined with *k*-means clustering loss (M2’s of 0.91 and 0.85 vs KG-DTC’s of 0.84 and 0.31). Thes advances are also visualized in the UMAP plots. The representation learned from the ablated model M3 without knowledge guidance has formed separated clusters. Still, the samples are gathered to form one giant cluster, failing to generate informative and well-separated clusters.

### Clustering reproducibility evaluation

After cross-validating our clustering model’s performance, we investigated the reproducibility of the clusters on external data. We applied the KG-DTC models trained with the discovery data (EXPEDITION1 + EXPEDITION3) to the external data (EXPEDITION2) and compared the clusters from each set. As a result, we identified similar clustering results from the external set (Table 1). The proportions of clusters were 48.1, 9.0, and 42.8% in discovery data and 43.0, 13.0 and 44.0% in external data. In addition, we found that applying the trained KG-DTC to the external set achieved the outcome classification accuracy (the of 0.84 and 0.32) and 0.33 silhouette scores of 3 clusters, which is similar to the results with the discovery set. In the UMAP plots, clustering results from the external set also achieved a compact and distinct shape. Our models show great representation quality and clustering reproducibility in the external set.

### Clustering stability evaluation

Now that we verified that the ORDTCR could reproduce similar clustering results with external data, we investigated whether the ORDTCR is stable enough to generate similar clustering results consistently. We evaluated our clustering model’s assignment stability using the leave-one-out (LOO) approach^5^ (Method: Clustering evaluation). We set leave-out sample size. *n* ∈ {1:50} Supplementary Fig. 1 showed that the average adjusted Rand Index (ARI) slightly fluctuated around 0.945 over different leave-out sample sizes. This finding demonstrated that our clustering model is stable, and the identified clusters are not statistical artifacts. Due to the stochastic nature of optimization, traditional clustering methods may produce inconsistent clustering results if the sample similarity is not well defined. The KG-DTC model projected the complicated multimodal longitudinal observation into a representation space where the sample similarity preserves the clinically meaningful patterns.

### Clustering interpretation and its statistical significance

After thoroughly validating our proposed clustering models, we investigated common patterns of patients within the clusters. To illustrate the within-cluster distribution of cognitive scores, brain regional volumes, and amyloid beta deposition, we plotted violin plots to visualize data distribution across three clusters (Fig. 3 (A)). In addition, the main drawback of neural-network-based clustering is its inability to explain how static data and longitudinal sequences map to the latent dimensions. To address this and obtain interpretable patterns, we calculated the feature importance scores to determine the important baseline and longitudinal variables for cluster assignment. We built a cluster assignment (i.e., S1, S2, and S3) classification model and adapted it to feature permutation attribution algorithm1 (Method: Clustering evaluation). The AUROC for cluster assignment classification tasks were 0.90, 0.94, and 0.96. Using this approach, we calculated the importance scores of individual variables (Fig. 3 (B)).

**Fig. 3.**
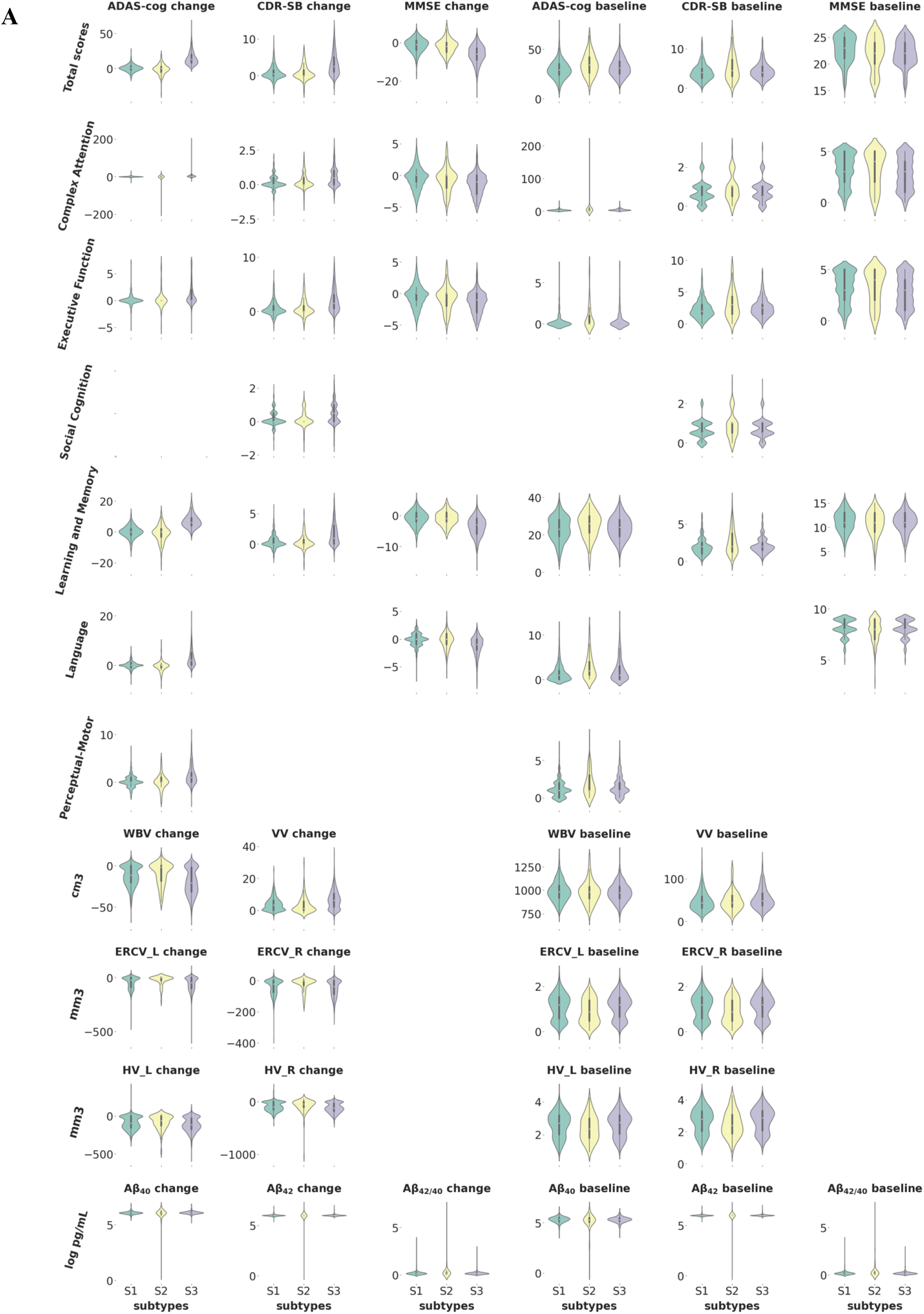
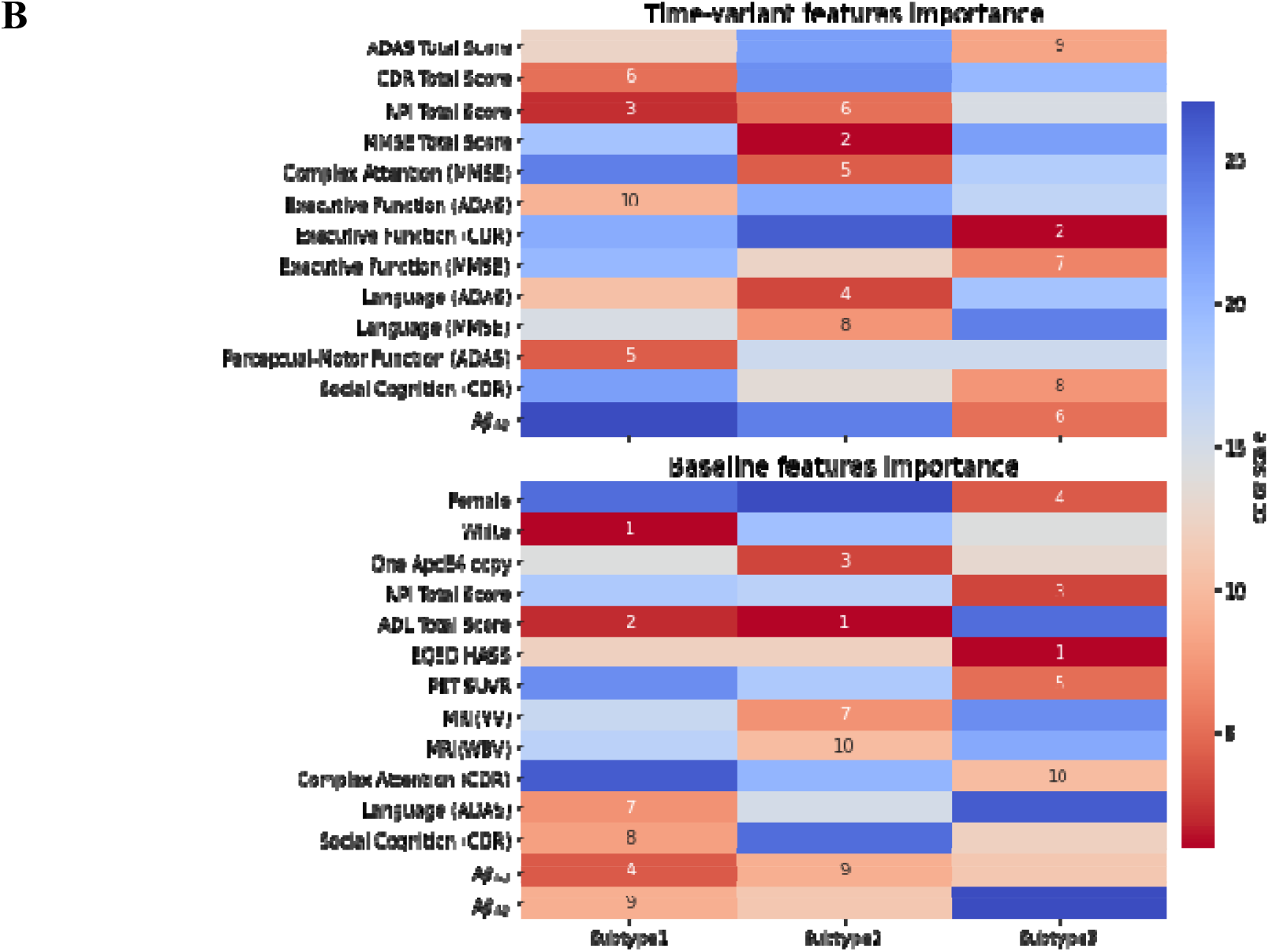
Feature distributions (A) and importance patterns (B) by clusters. Violin plots (A) illustrate the within-cluster distribution of cognitive scores, brain regional volumes, and amyloid beta deposition at baseline and change to the endpoint, respectively. Note the unequal sample size among the cluster types: S1 (N=741), S2 (N=139), and S3 (N=659). Heatmap (B) visualizes the top important baseline and time-invariant features.

To provide more detailed characterizations of each subtype, we statistically tested the important individual variables to determine whether they were distributed differently across clusters. As a result, we identified important features that distinguish the cluster assignment and generated profiles for each cluster. The profiles of clusters (or AD subtypes) have statistically distinctive characteristics. Also, the clusters were closely associated with the primary clinical outcomes of the trials. We identified three AD subtypes (Subtype 1 (S1), Subtype 2 (S2), and Subtype 3 (S3)) with distinct patterns in impaired cognitive function (C), neurodegeneration (N), and amyloid beta (A) (Table 2), as well as clinical outcomes in the trials (Fig. 4).

**Table 2.**
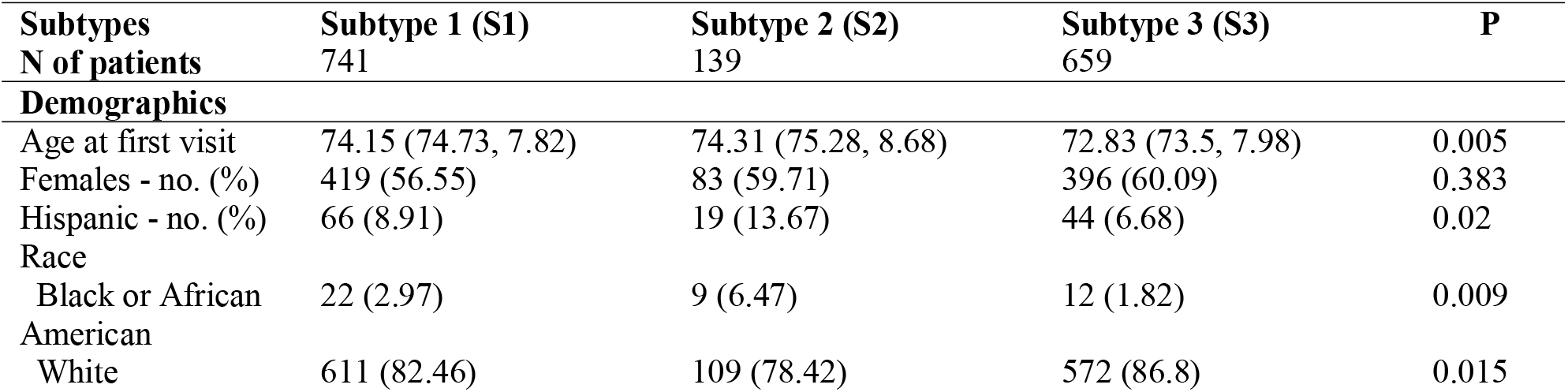
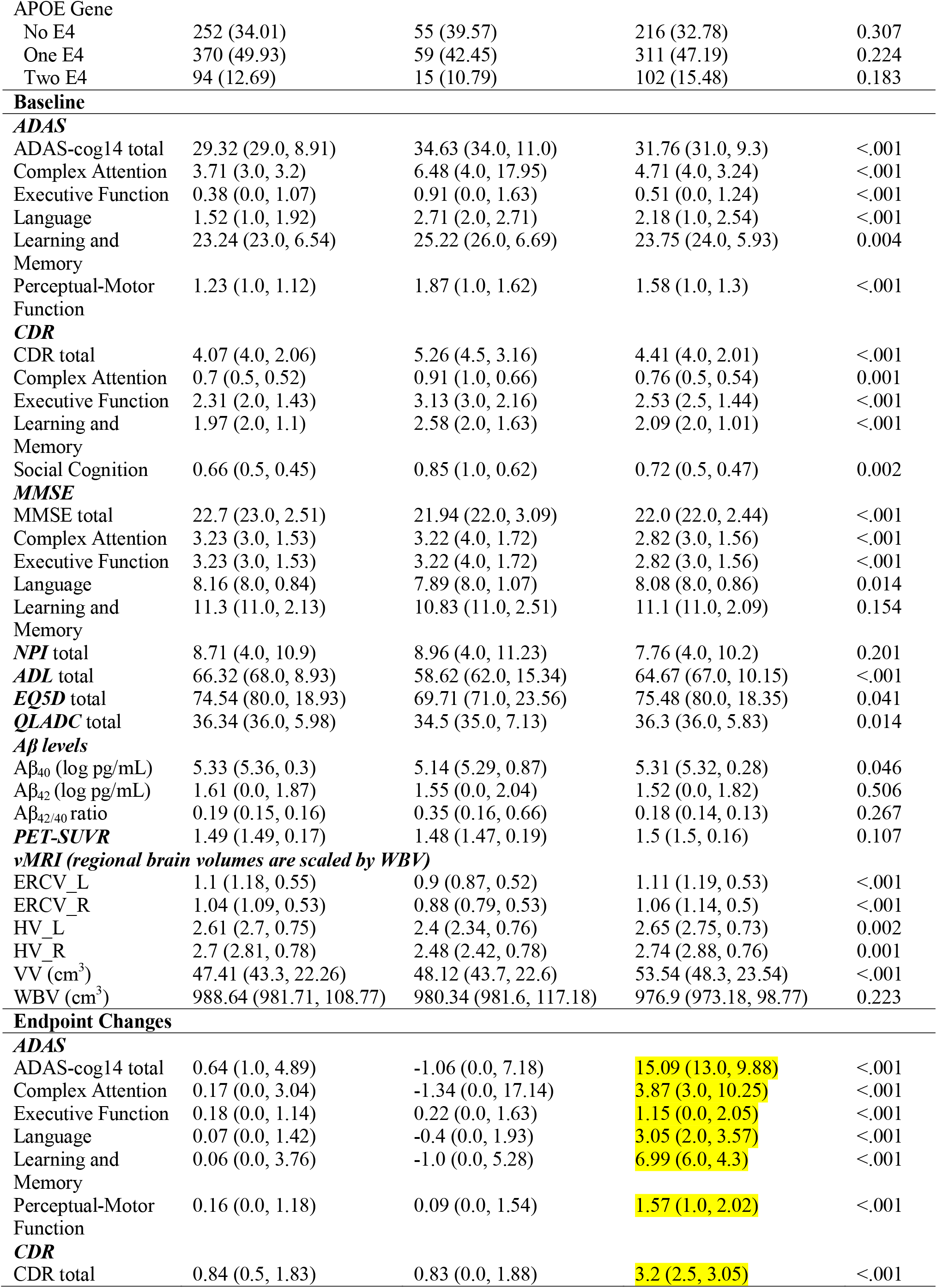
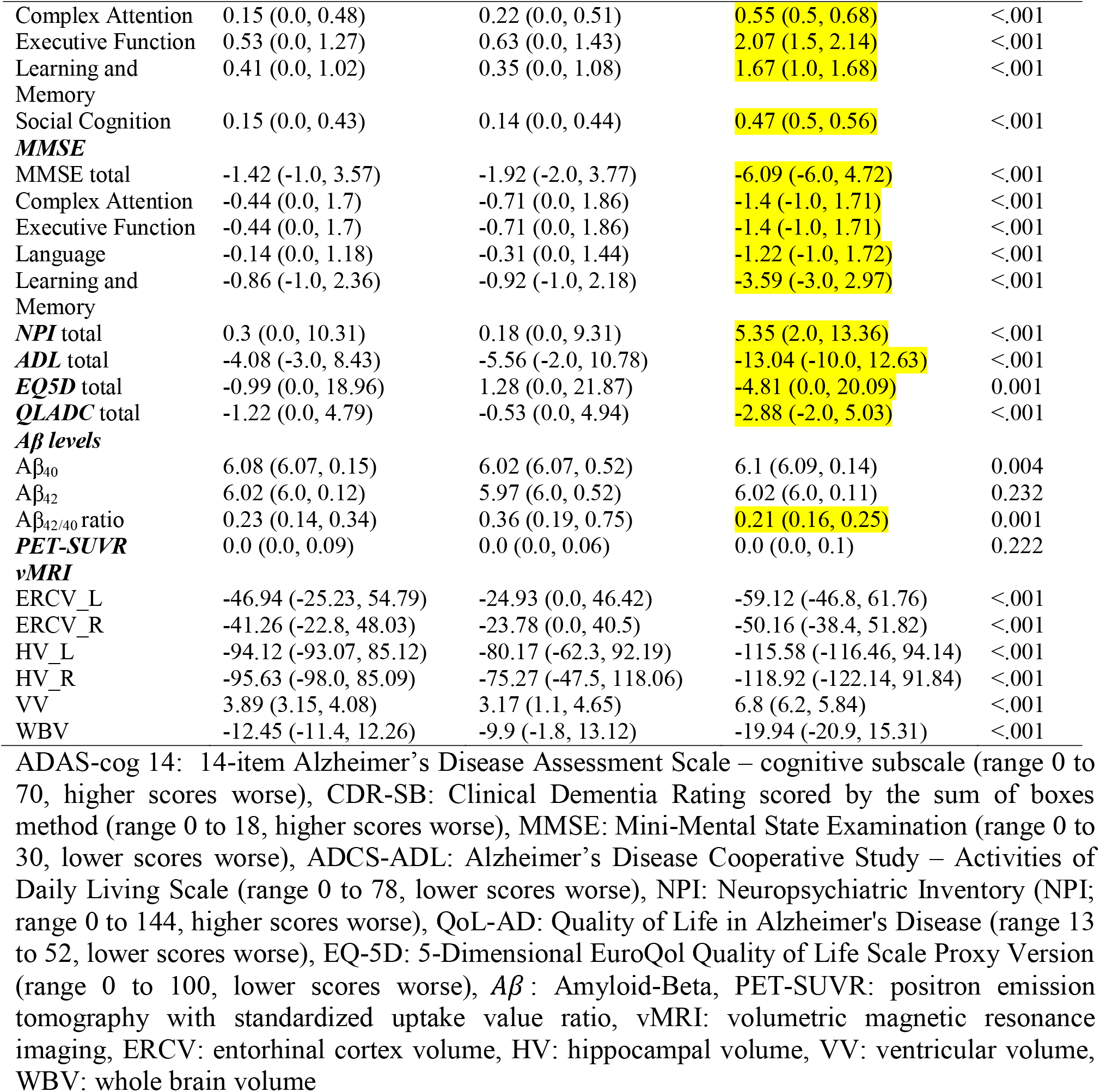
Characteristics of Subtypes at Baseline and Endpoint Changes. (The cells represent mean (median, std) unless specific illustration)

**Fig 4.**
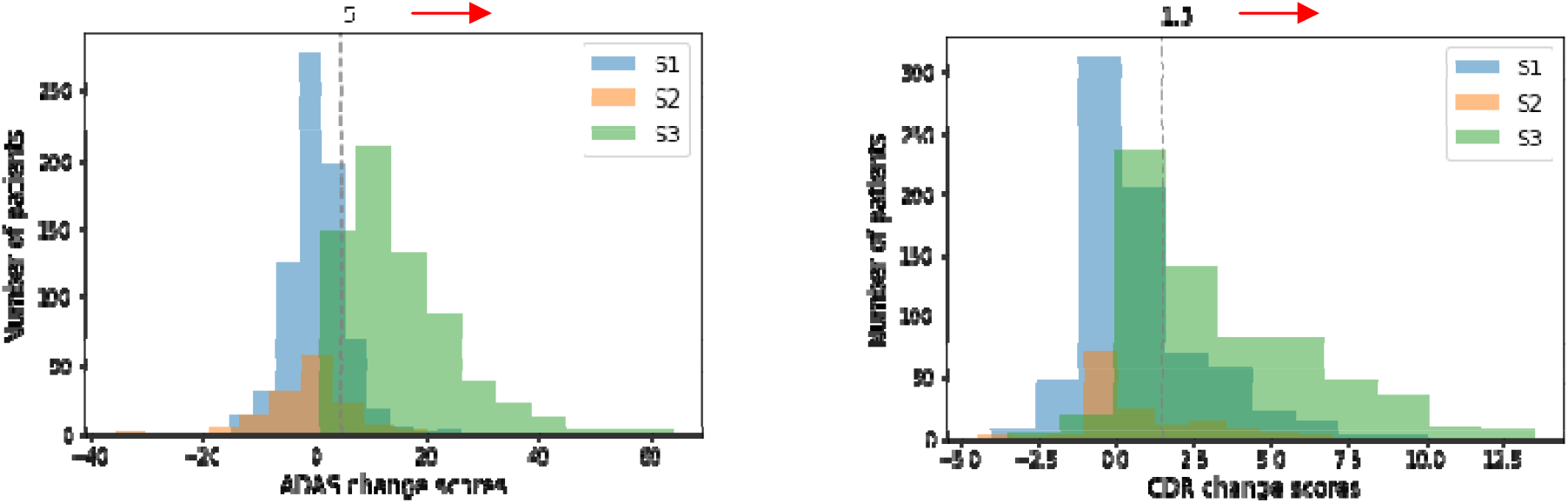
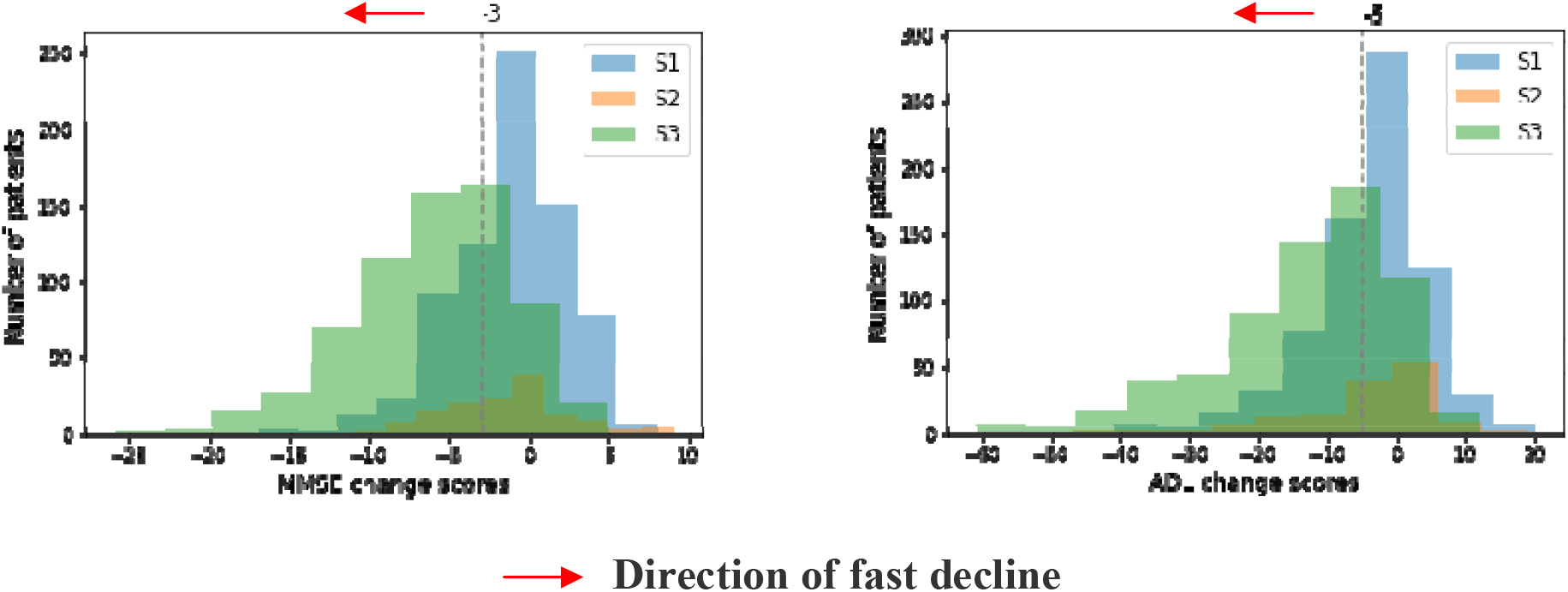
Distribution of different clinical outcome variables for identified clusters.

**Fig 5.**
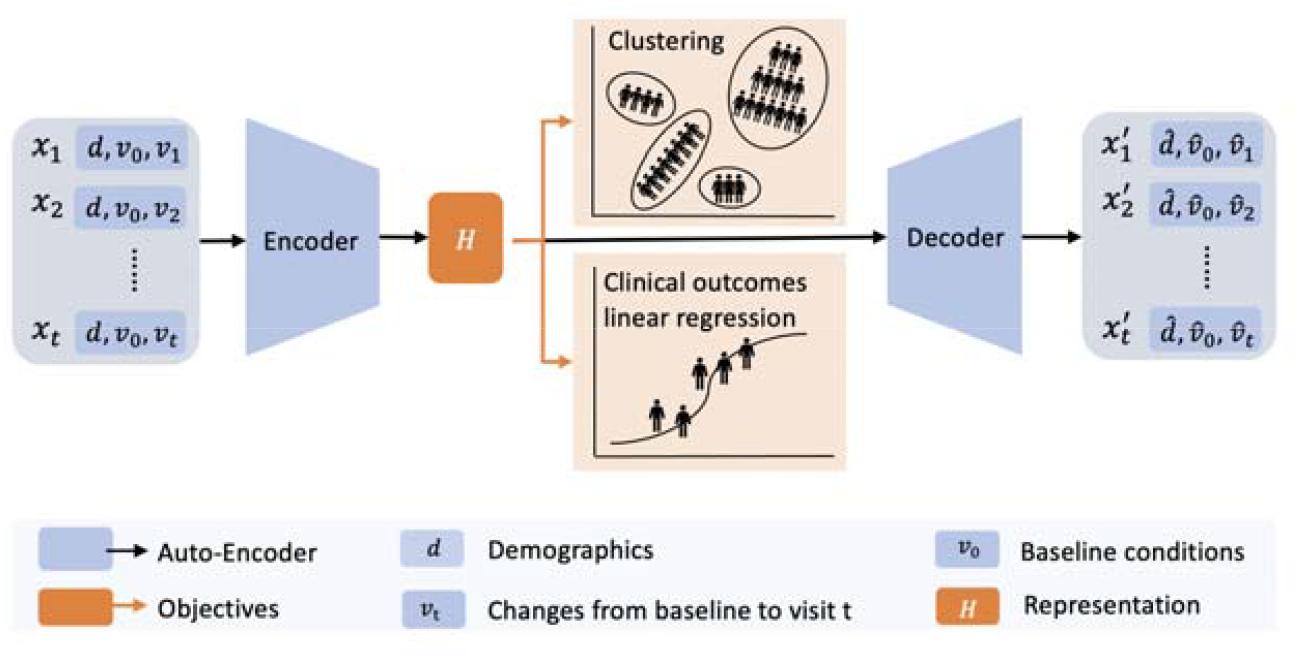
KG-DTC model structure. KG-DTC architecture. We used a temporal autoencoder to encapsulate the multivariate baseline (,) and temporal features () into a hidden representation *H*. The hidden representation *H* is jointly derived to minimize the clustering loss and clinical outcome regression loss.

Specifically, S1 (48.1% in EXPEDITION1+EXPEDITION3; 43.0% in EXPEDITION2) had the largest cohort with 741 Alzheimer spectrum patients. S2 (9.0% in EXPEDITION1+EXPEDITION3; 13.0% in EXPEDITION2) had the smallest cohort with 139 patients containing the highest proportion of the Hispanic or Latino population (8.91% in S1 vs. 13.67% in S2 vs. 6.68% in S3, p=0.02; We omitted S1, S2, and S3 below). S2 also had the highest proportion of the Black or African American population (2.97% vs. 6.47% vs. 1.82%, p=0.009) and the lowest proportion of the White population (82.46% vs. 78.42% vs. 86.8%, p=0.015). S3 (42.8% in EXPEDITION1+EXPEDITION3; 44.0% in EXPEDITION2) had 659 slightly younger patients on average (74.15 vs. 74.31 vs. 72.83, p=0.005).

At baseline, S1 and S3 showed similar overall shapes of data distribution in the violin plots. The APOE genotype didn’t show significant differences between subtypes. However, S2 had the highest portion of patients without the APOE 4 gene and the lowest portion of APOE 4 carriers. In terms of baseline cognitive assessments, S2 consistently exhibited higher scores across multiple domains and total scores of ADAS and CDR than S1 and S3 (e.g., 29.32 vs. 34.53 vs. 31.76 of average ADAS total score, p<.001; 4.07 vs. 5.26 vs. 4.41 of average CDR total scores, p<.001), which indicated worse cognitive performance or more severe disease symptoms. On the MMSE scale (a measure of cognitive function where lower scores indicated more severe cognitive impairment), S2 had a slightly lower mean score compared to S1 and S3 (e.g., 22.7 vs. 21.94 vs. 22.0 of average MMSE total scores, p<.001). S1 consistently exhibited the most minor cognitive impairment (see details in Table 2). For levels, S2 had lower (5.33 vs. 5.14 vs. 5.31 log pg/mL, p=0.046), though the differences across the subtypes were significant only for, not for or the ratio. Furthermore, neuroimaging data suggested that patients in S3 generally showed greater entorhinal cortex and hippocampal volumes than S1 and S2, while S2 showed the lowest brain regional volumes (see details in Table 2). S3 also retained the most significant average ventricular volume (47.41 vs. 48.12 vs. 53.54 cm^3^, p<.001). The PET-SUVR and whole brain volume didn’t show significant differences across subtypes.

We then evaluated longitudinal trends using endpoint changes for time-variant variables. S1 and S2 showed similar shapes of cognitive data distribution in violin plots, while S1 and S3 showed similar shapes of neuroimaging data and levels. We observed that S3 significantly increased in scores of multiple domains and total scores of ADAS and CDR (e.g., 0.64 vs. −1.06 vs. 15.09 of average ADAS total change score, p<0.001; 0.84 vs. 0.83 vs. 3.2 of average CDR total change scores, p<.001), indicating a drastic cognitive decline. Interestingly, S2 showed preserved cognitive domains of complex attention (−1.34), language (−0.4), and learning and memory (−1.0) measured by ADAS. On the MMSE scale, S3 showed significantly decreased scores (e.g., −1.42 vs. −1.92 vs. −6.09 of average MMSE total change scores, p<.001), indicating deterioration. In levels, S3 showed the most increase in (6.08 vs. 6.02 vs. 6.1 log pg/mL, p=0.004) and the least increase in the ratio (0.23 vs. 0.36 vs. 0.21, p=0.001), while S2 was the opposite. In terms of brain imaging changes, S3 showed the most significant loss in regional brain volume (e.g., −46.94 vs. −24.93 vs. −59.12 mm^3^ of average left entorhinal cortex change volume, p<.001; −94.12 vs. −90.17 vs. −115.58 mm^3^ of average left hippocampal change volume, p<.001) and whole brain volume (−12.45 vs. −9.9 vs. −19.94 cm^3^, p<.001), which indicated brain atrophy that might correlate with the cognitive decline observed in cognitive measurements. In addition, S3 showed the most significant increase in ventricular volume (3.89 vs. 3.17 vs. 6.8 cm^3^, p<.001), suggesting greater brain atrophy. Here, S2 always showed the most minor changes compared to S3.

Finally, we plotted the distribution of four clinical outcome measures (i.e., ADAS, CDR, MMSE, and ADL) across clusters. We separated the population into different paths of progression using the middle value of each clinical outcome. We observed that the distribution of S3 lay mostly in the fast decline directions across all four measures.

## Discussion and Conclusions

Our objective was to shed light on the heterogeneity of Alzheimer’s Disease progression patterns by developing a cutting-edge, deep-learning framework to identify AD subtypes with static and longitudinal features. Specifically, we developed a representation model called the knowledge-guided deep temporal clustering representation (KG-DTC) to generate cluster-friendly and clinical outcome-related representations. The learned representation was examined with effectiveness in discriminating the heterogeneity of dynamic and complex disorders in th AD cohort. By applying this approach to the pooled randomized clinical trial data, we identified three distinct AD subtypes: S2 had the highest cognitive impairment at the baseline but preserved progression at the endpoint, while S3 had significant deterioration at the endpoint. S1 represented those who have milder symptoms at baseline and often demonstrate less decline over time. This finding provided a novel understanding of AD progression in combination with knowledge of neuropathological and clinical heterogeneity, which may pave the way for individualized AD progression forecasts and customized treatments for specific AD subtypes.

We employed a seq2seq architecture to encode multivaria longitudinal data and integrated *k*-means clustering loss and clinical outcome classification loss to form clinically meaningful cluster structures. Here, representation learning aimed to map the high-dimensional complex data to lower-dimensional space. However, traditional representation learning methods are not designed for clustering tasks. To enable the learned representations to favor clustering patients, we integrated the *k*-means clustering loss to encourage the representations to form clusters. Additionally, previous studies mostly focused on identifying AD subtypes in a purely unsupervised way^6,7,15^. While these studies obtained the AD subtypes without considering long-term clinical outcomes, the identified AD subtypes may not reflect our existing knowledge of outcomes of clinical interest. We filled this gap by introducing a clinical outcome supervision task; we employed a classification loss in the model to enable the learned representations to be informative with multiple clinical outcomes and unveil clinically meaningful and actionable insights.

We identified three distinct AD subtypes from pooled randomized clinical trials, which demonstrated heterogeneity in the presentation and progression of Alzheimer’s disease. These subtypes were aligned with the NIA-AA’s AT(N)(C) classification (based on fluid biomarkers, neurodegeneration, and cognitive symptoms) and with rapid/slow progressors (based on cognitive presentation) ^16,17^. At the baseline, S2 has a higher representation of Hispanic and Black or African American populations. Race and ethnicity variations among the subtypes could hint at potential genetic or environmental risk factors specific to different populations.^18,19^ Clinically, S2 manifests a more severe cognitive deficit at baseline, as evidenced by elevated ADAS and CDR scores, reduced MMSE scores, and lower levels of brain regional volumes. These suggested that S2 might represent a more severe or rapidly progressing form of the disease. However, when it comes to endpoint changes, S3 appears to deteriorate at a faster rate than S2 regarding cognitive measures and neurodegeneration, indicating that Subtype 3 might experience a delayed but rapid cognitive decline, which suggested an A(−) N(+) C(+) profile in the NIA-AA framework. Moreover, the significantly greater increase versus other subtypes of *Aβ*_42/40_ in S2 suggests potential variations in amyloid pathology or deposition. Interestingly, certain cognitive domains in S2 showed lesser deterioration or even improvement, coupled with a reduced rate of regional brain atrophy, which suggested an A(+) N(−) C(−) profile in the NIA-AA framework.

Our data-driven subtyping can contribute to connecting the subtypes into the primary endpoint of clinical trials, which will facilitate patient specific therapeutic development. Traditional clinical trials often treat Alzheimer’s patients as a homogenous group. However, the existence of distinct subtypes suggests that treatments could be more effective if they were more tailored. Targeted interventions can be designed by identifying which subtype an individual belongs to, which may increase the likelihood of therapeutic success. Moreover, aligning data-driven subtypes with established biomarker frameworks, such as the NIA-AA’s AT(N)(C) classification, allows for a more integrated understanding of the disease. This can help in the identification of novel biomarkers and the development of therapies targeting specific pathways.

Our study has some limitations. We did not include the comorbidity risk factors when modeled in the context of AD heterogeneity. This reduced the interpretation ability of identified AD progression patterns that may be affected by other diseases or drugs. In the future, we envision combining and comparing comorbidities longitudinally, thus extending our current analyses to understand the contribution of comorbidities in AD subtypes. The inclusion of patients from two different trials is an advance since it increased the variability in the sample and therefore, represented the AD population better, but it is also a limitation due to data variability in multiple studies considering different efficacy measure standards and irregular visit intervals. Future work can focus on better visit alignment and missing imputations. Our model has to pre-specify the number of clusters in *k*-means clustering loss. However, we do not exactly know how many subtypes are in the datasets naturally, and it is sensitive to which datasets are used. Future studies should design a structure that can automatically optimize the selection of the best N of clusters during the model training process.

In conclusion, we discovered three longitudinal patterns of AD subtypes by a novel outcome-regularized deep temporal clustering approach. Our study is an important step towards solving an unmet need, i.e., uncovering the subtypes of AD disease progressions with observed heterogeneity in neurology and biology. Moreover, our proposed models unravel the heterogeneity in AD that can enable precision medicine and potentially lead to successful disease-modifying treatments in the future.

## Methods

### Study design and dataset

Figure 1 shows the overall study design, including data preprocessing, subtype discovery, model training, and external validation, and clustering significance and stability evaluation. In this retrospective study, we used patient data in placebo groups from three randomized AD trials: 505 in EXPEDITION1 (NCT00905372^13^, 2009-2012), 518 in EXPEDITION2 (NCT00904683^13^, 2009-2012), and 1064 in EXPEDITION3 (NCT01900665^14^, 2013-2017). Patients in placebo groups allow us to investigate the disease progression without being confounded by the exposure to experimental drugs. The three trials tested Solanezumab, a humanized anti-amyloid monoclonal antibody, for its efficacy in slowing AD decline. They had similar entry criteria, including patients 55 years or older who met the criteria of the National Institute of Neurological and Communicative Diseases and Stroke–Alzheimer’s Disease and Related Disorders Association for probable AD. EXPEDITION1 and EXPEDITION2 included mild to moderate patients with MMSE scores of 16 to 26, while EXPEDITION3 included mild patients with MMSE scores of 20 to 26. They then underwent 80-week observations. For each patient, the time interval between two adjacent longitudinal measurements ranges from 4 to 16 weeks.

### Variables

We included 8 demographics, 28 baseline clinical conditions, and 28 longitudinal clinical conditions’ changes in our model. The clinical conditions included cognitive assessments, imaging biomarkers, fluid biomarkers, quality-of-life assessments, and a neuropsychiatric assessment from three trials. The cognitive assessments included ADAS-cog14 (range 0 to 90, higher scores worse), ADCS-ADL (range 0 to 78, lower scores worse), CDR-SB (range 0 to 18, higher scores worse), and MMSE (range 0 to 30, lower scores worse). Imaging biomarkers contained v-MRI volumes of the whole brain and regional brains (e.g., hippocampi, entorhinal cortices, and ventricles) and amyloid PET imaging for the composite summary standard uptake value ratios (SUVRs). The fluid biomarkers are plasma *Aβ* levels. The quality-of-life assessments are QoL-AD (range 13 to 52, lower scores worse) and EQ-5D Proxy (range 0 to 100, lower scores worse). The neuropsychiatric assessment included NPI (range 0 to 144, higher scores worse). Supplementary Table 1 provides a summary of all variables in each cohort.

### Data Preprocessing

We preprocessed the datasets by applying data harmonization, missing value imputation, and data transformation to all three trials. As each study can have variables with different names or units, careful harmonization was conducted. We matched the variable by reviewing the Case Report Form in each trial. We selected visits with assessments conducted or biomarkers collected (i.e., visits 1, 2, 3, 4, 5, 6, 10, 13, 16, 19, and 23.0 for EXPEDITION 1 and 2; visits 1, 2, 3, 5, 9, 12, 15, 18, and 22 for EXPEDITION 3). For features that have longitudinal changes, we calculated change values from the baseline to the visits, thus patients’ longitudinal trends were represented as a set of change value sequences. To deal with missing values, we leveraged various imputation strategies. Within a sequence, a missing value indicated that a test was not conducted during that visit. Assuming a patient’s condition stayed stable before and after a test visit, we filled in missing values by propagating the existing values forward and backward along a sequence. The remaining missing values were imputed by chained equations (MICE), which is a multiple imputation method that builds a multivariate predictive model to infer the missing values using the remaining features in a round-robin fashion^20^. Data from different distributions were carefully standardized. We log-transformed skewed distributions (i.e., levels) into Gaussian and Gaussian into z-distributions to make variables comparable. To reduce cognitive variable redundancy, specific cognitive test items (e.g., test of Comprehension of Spoken Language, Word-Finding Difficulty, and Naming Objects and Fingers in ADAS) were grouped into cognitive functions as grouped variables (e.g., language in ADAS). Overall, the cognitive test items of ADAS, CDR, and MMSE were grouped into 6 cognitive functions (i.e., complex attention, executive function, language, learning and memory, perceptual-motor function, and social cognition), respectively.

### Subtype discovery: Knowledge-Guided Deep Temporal Clustering (KG-DTC) model

We developed a novel knowledge-guided deep temporal clustering model (see Fig. X). that identifies patient clusters by deriving cluster-friendly embedding of temporal observations in an end-to-end framework. Our model is built upon a sequence-to-sequence structure (seq2seq) that encapsulates time-invariant and time-variant observations into a representation. Here, a technical challenge is that the representation from the seq2seq does not necessarily form a cluster. Motivated by prior research that embeds clustering into representation learning^21,22^, we encouraged the representation to form clusters by incorporating the *k*-means clustering loss during training. Clinical outcomes are the main results that are measured at the end of a study to see whether a given treatment worked. To leverage clinical outcomes as knowledge, we also guided the representation to be discriminative to clinical outcomes of interest, in order to identify highly responsive groups and less responsive groups. Details of each component are as follows.

### Temporal autoencoder (Seq2seq)

We used gated recurrent units (GRUs) autoencoders (AEs) for the seq2seq structure. We first instantiated the encoder as a batch normalization layer connected with a 2-layer stacked GRUs^23^ to capture temporal and multiscale characteristics of input data. We then utilized a single-layer GRU as the decoder to reconstruct the input. GRU is a variation of RNN with an additional relevance gate and updated gate, which can capture time dependencies over different periods and thus efficiently handles temporal patterns. AEs learn the embedded representations for high-dimensional data by reconstructing the input series and minimizing the reconstruction loss.

Given *N* patients, the patients data were divided into three parts: demographic information 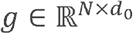, baseline clinical conditions 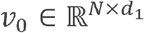, and longitudinal clinical condition changes over visit t 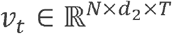, where t ∈ {1,…, *T*}and *T* was the total number of visits, and *d*_0_,*d*_1_,*d*_2_ were the number of features in each part, respectively. The static components *g* and *v*_0_ were repeated for each time step *t*. We then concatenated the *g, v*_0,_ *and v*_*t*_ to form a sequence of feature vectors for patient *i* at visit *t*, denoted as *x*_*i,t*_ = [*g*; *v*_0_; *v*_*t*_;], *x*_*i,t*_ ∈ ℝ ^*D*^ where *D* = *d*_0_ + *d*_1_ + *d*_2_, was the total number of features. Therefore, the sequence of concatenated vectors for patient *i* across all visits can be represented as: *x*_*i*_ ∈ ℝ ^*T*×*D*^. We drop *i* for simplicity in the following notation.

We first applied batch normalization over input *x* ∈ ℝ ^*T*×*D*^ The output *y* ∈ ℝ ^*T*×*D*^ is

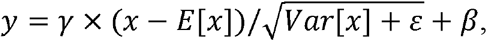

where *E*[*x*] is the expectation of *x, Var*[*x*]is variance of *x*, γ and *β* are parameters to be learned, and ε is a constant added for numerical stability. For simplicity, we still use *x* as the output of batch normalization in the following notation. We then applied multi-layer GRU RNN to the *x* as encoder. Given input 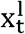 at visit t, each layer l computed the following functions:

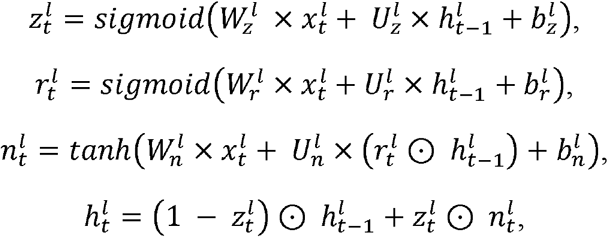

where 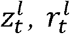, and 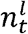 were update, reset, new gates, respectively. The update gate 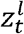 helped the model to determine how much of the past information needs to be passed to the future. The reset gate 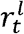 defined how much of the past information to forget. The new gate combined the new input with the past hidden state to create new candidates for the hidden state. The *W* and *U* were the weight matrices and *b* was the bias for each gate in layer *l*. The 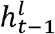 was the past hidden state from the previous visit in the same layer. The 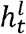 was the combination of the new gate and the past hidden state, controlled by the update gate. The output 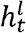 was used as the input 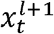 for the next layer in the stacked GRU. Therefore, the final hidden representation from the encoder was 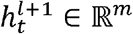, where *m* was the hidden size.

The single-layer GRU decoder is essentially another RNN layer that takes hidden representation 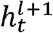 generated by the encoder and aims to reconstruct the original input sequence. The computation process is the same as the encoder and the final hidden state returned by the decoder is 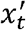. This sequence 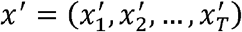 is the reconstructed version of the original input sequence. We used mean square error (MSE) to evaluate the quality of the reconstruction, which can be formulated as

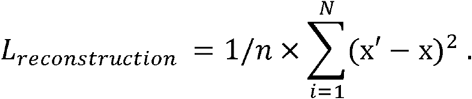

### *K*-means clustering loss

We denoted the hidden representation 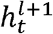 for *N* patients as *H* ∈ ℝ ^*N*×*m*^. It is noted that the *H* obtained from the temporal autoencoder does not guarantee distinctive clusters. Hence, following previous work^22^, we encouraged *H* to form clusters while maintaining the reconstruction by regularizing the AEs through a soft *k*-means objective, defined as follows

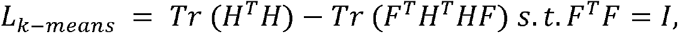

where *F* ∈ ℝ ^*N*×*k*^ denoted the cluster membership matrix, *F*^*T*^ *F* = *I*, and *k* was the number of clusters. Minimizing the *k*-means clustering loss with *H* was equivalent to maximizing the trace *Tr* (*F*^*T*^ *H*^*T*^ *HF*)^23^. Since the learning of *H* was dynamic instead of static, the training process consisted of iteratively updating *F* and *H*. When fixing *F*, updating *H* can follow the standard stochastic gradient descent (SGD), encouraging the representations to form cluster structures. While fixing *H*, according to the Ky Fan theorem^24^, we updated *F* using the closed-form solution to the trace maximization problem by computing the k-truncated singular value decomposition (SVD) of *H*. We fixed a cluster count of four in *k*-means clustering loss, which was commonly used in multiple previous AD subtyping studies^25–27^.

### Knowledge guidance

To enable the learned representation to be discriminative to the clinical outcomes, we introduced a multi-target multi-linear regression to encourage the learned representations to predict multiple clinical outcomes. We selected two primary clinical outcome assessments (i.e., total change of CDR-SB and ADAS-cog14 from baseline to end of observation) in the trials. We jointly trained the encoder that can detect multiple continuous clinical outcomes. Each patient had two clinical outcomes *y* ={*y*_*CDR*_, *y*_*ADAS*_}. We predicted the clinical outcomes ŷ_*j*_ *=W*_*fc*_ × *H +b*, where *W*_*fc*_ ∈ ℝ ^2×*m*^ were the weights of the fully connected layers and *b* is the residual term. The loss between ground truth and predicted results are defined as

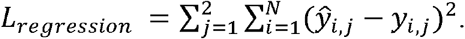

We jointly optimized the three training losses *L*_*reconstruction*_, *L*_*k-means*_, and *L*_*egression*_. To mitigate detrimental gradient interference in multi-objective learning, we use the gradient surgery to project conflicting gradients into the norm of other gradients^28^. Gradient surgery is a technique used to separate the gradients of different tasks, allowing the model to learn each task independently. This can help to prevent the gradients of one task from interfering with the gradients of another task, which can lead to suboptimal performance on one or more tasks. Throughout the training process, the batch size and hidden state feature size were set to 64 and 32, respectively.

After we trained the model, we obtained the deep temporal representation *H* for patients, which was used for downstream clustering tasks to determine distinct AD subtypes. Although the *k*-means clustering in our model provides the cluster membership (i.e., *F*), we trained a Gaussian mixture model (GMM) and generated final clusters to account for soft membership. To determine the number of clusters k, we calculated the silhouette scores for k from 2 to 10 (Fig. 6 (A)). The silhouette score measures the similarity of an object to its own cluster compared to other clusters. It ranges from −1 to 1, with a higher value indicating more cluster separation. GMM is soft clustering, thus difficult to assign one sample to one cluster. We address this challenge by setting the probability threshold of membership. The distribution of cluster membership probabilities shows that subjects are concentrated in the probability interval of more than 0.9 (Fig. 6 (B)). Thus, after obtaining the clusters from GMM, we selected the representative “insider” patients for each cluster who had a cluster membership probability larger than 0.9.

**Fig 6.**
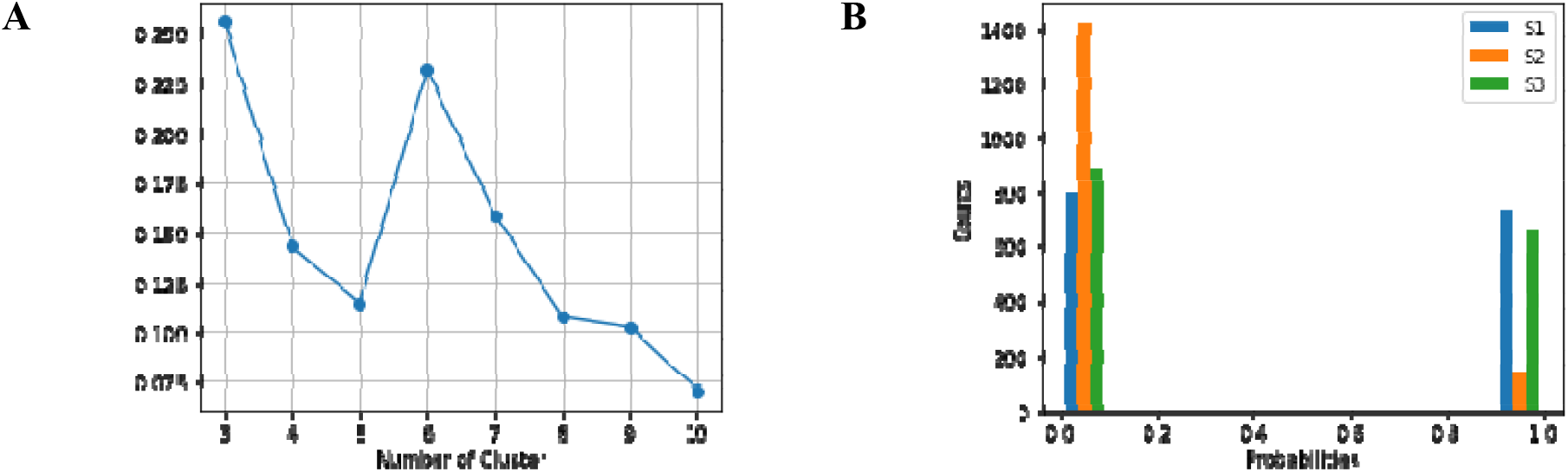
Distribution of silhouette scores and predicted membership probabilities.

### Model evaluation

#### Cross-validation

We cross-validated the model on the discovery set to find the best model (as illustrated in Fig. X.). To this end, we randomly partitioned the discovery set into training, validation, and test sets in a ratio of 8:1:1 for training, best model selection, and final model performance testing. In the cross-validation we focused on balancing the tradeoff between multiple objectives: outcome prediction accuracy and clustering performance. We evaluated the model’s capability to learn a representation that could effectively predict clinical outcomes. Despite incorporating clinical outcomes as a knowledge regularizer during training, the representation might not always predict these outcomes accurately due to the optimization of clustering.

The performance of the regression tasks was measured by the values. Higher values indicate that the model accounts for a good amount of variance of clinical outcome variables. We also evaluated our model’s clustering performance by silhouette scores and UMAP visualization. A higher silhouette score indicates betterer clustering ability, and UMAP facilitates the mapping of learned representations into a 2D plot to visualize the shape and distribution of clusters.

#### Ablation study

To examine the contributions of individual components (i.e., clustering optimization and knowledge infusion) within our full model, we compared the full model against three ablated models to assess the importance of each module: M1 with only temporal encoder, M2 with temporal encoder and *k*-means clustering optimization, M3 with temporal encoder and knowledge infusion regression tasks. Performance is also assessed by silhouette score, UMAP plots, and clinical outcome prediction accuracy.

### Clustering evaluation

#### Reproducibility

To further demonstrate the generalizability of the model and reproducibility of the clustering patterns, we externally validated the trained model in an independent cohort. We applied the trained KG-DTC model using the discovery set to the external set and compared the clusters derived from both sets. We compared the distribution of clustering membership by comparing UMAP plots. To evaluate the reproducibility of clinical implication (i.e., clinical outcome prediction) and clustering performance, we calculated the prediction accuracy and silhouette scores on the external set.

#### Stability

We quantified the clustering stability using the ARI. ARI computes the similarity between two clustering results by counting pairs that are assigned to the same clusters in both clustering results^29^. It ranges from −1 to 1, with a score of 1 indicating a perfect match between two clustering results. To investigate the stability of clustering assignment against modest data perturbations, we adopted a LOO approach. In this approach, we randomly left out a small number of patients *n* ∈ {1:50}in the external set, applied the trained KG-DTC model, and obtained perturbed clustering results. This process was repeated 200 times for each setting, which allowed us to compare the stability of clustering. We measured the similarity between the 200 different clustering results by calculating the ARI score.

#### Interpretation

We conducted a feature importance analysis using a permutation-based approach to identify variables that uniquely determine the clusters. We first built classification models to predict cluster membership for patients by adding two fully connected layers to the trained KG-DTC model. To measure the importance of each feature (including both time-variant and time-invariant features) in determining the cluster membership, we calculated feature importance scores by permuting the inputs of the model. Specifically, we randomly shuffled the values patient-wise for both static and time-varying features and then measured the changes in the performance of the model on this shuffled set. Features with a larger decrease in performance were more important in determining the clusters.

To gain a deeper understanding of clustering, we plotted the distribution of four clinical outcome measures (i.e., ADAS, CDR, MMSE, and ADL) across clusters. By separating the population into different paths of progression using the middle value of each clinical outcome, we were able to infer which cluster characteristics were informative regarding progression speed.

#### Statistical significance tests

We conducted post-hoc statistical tests to identify variables that showed significant differences across clusters. For non-parametric variables, we used the Kruskal-Wallis test. For variables that followed a Gaussian distribution, we employed the ANOVA test, and for categorical variables, we utilized the Chi-squared test. These statistical analyses enabled us to determine whether baseline clinical conditions and clinical condition changes at the final visit were significantly different across clusters. This provided valuable insights into the distinct factors contributing to the separate clusters identified by our model.

## Data Availability

All data produced in the present study are available upon reasonable request to the authors

## Author contributions

Concept and design: DW, XM, PS, XJ, and YK. Data analysis: DW, XM, XJ, and YK. Literature review: DW, PS, XJ, and YK. Model development: DW, XM, XJ, and YK. Verification of results: DW, XM, PS, XJ, and YK. Strategic guidance and oversight: PS, XJ, and YK. All authors had full access to all the data, contributed to data interpretation, drafted and edited the manuscript, approved the final manuscript, and accepted the responsibility to submit it for publication.

## Acknowledgement

The authors would like to thank Christine M. Farrell, Ph.D. for her contribution to securing database access for this study, and Kristofer Harris for his contribution to clarifing the importance of identifying fast progressors without explicit definitions. This publication is based on research using data from data contributors Eli Lilly that has been made available through Vivli, Inc. Vivli has not contributed to or approved, and Vivli and Eli Lilly are not in any way responsible for, the contents of this publication.

## Data availability

Data access restrictions apply. Interested parties may contact Eli Lilly and Company for dataset licensing.

## Appendix Supplementary Materials

**Supplementary Figure 1.**
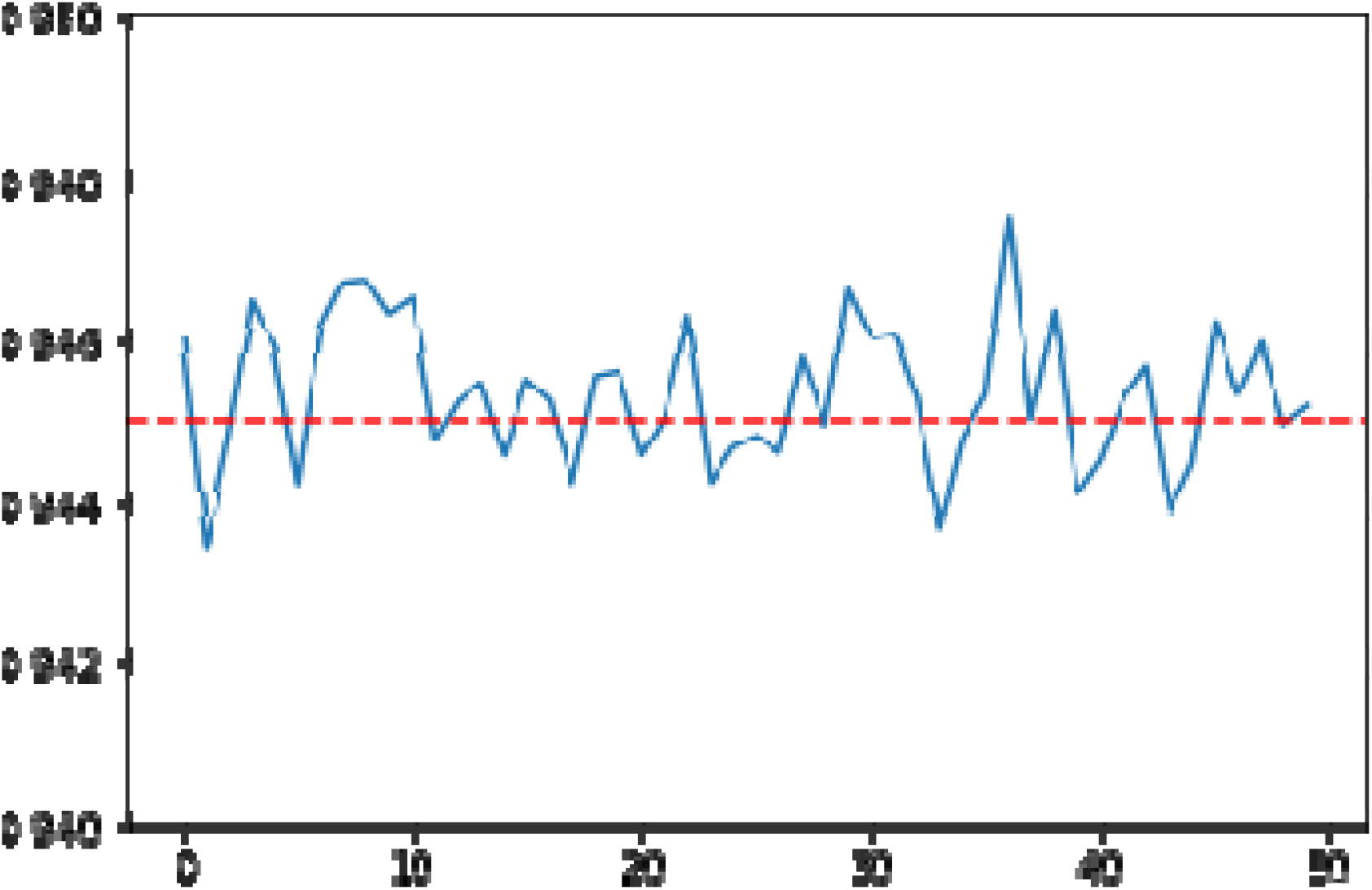

